# Performance of African-ancestry-specific polygenic hazard score varies according to local ancestry in 8q24

**DOI:** 10.1101/2021.01.20.21249985

**Authors:** Roshan A. Karunamuni, Minh-Phuong Huynh-Le, Chun C. Fan, Wesley Thompson, Asona Lui, Maria Elena Martinez, Brent S. Rose, Brandon Mahal, Rosalind A. Eeles, Zsofia Kote-Jarai, Kenneth Muir, Artitaya Lophatananon, UKGPCS collaborators, Catherine M. Tangen, Phyllis J. Goodman, Ian M. Thompson, William J. Blot, Wei Zheng, Adam S. Kibel, Bettina F. Drake, Olivier Cussenot, Géraldine Cancel-Tassin, Florence Menegaux, Thérèse Truong, Jong Y. Park, Hui-Yi Lin, Jack A. Taylor, Jeannette T. Bensen, James L. Mohler, Elizabeth T.H. Fontham, Luc Multigner, Pascal Blanchet, Laurent Brureau, Marc Romana, Robin J. Leach, Esther M. John, Jay H. Fowke, William S. Bush, Melinda C. Aldrich, Dana C. Crawford, Jennifer Cullen, Gyorgy Petrovics, Marie-Élise Parent, Jennifer J. Hu, Maureen Sanderson, Ian G. Mills, Ole A. Andreassen, Anders M. Dale, Tyler M. Seibert, The PRACTICAL Consortium

## Abstract

We previously developed an African-ancestry-specific polygenic hazard score (PHS46+African) that substantially improved prostate cancer risk stratification in men with African ancestry. The model consists of 46 SNPs identified in Europeans and 3 SNPs from 8q24 shown to improve model performance in Africans. Herein, we used principal component (PC) analysis to uncover subpopulations of men with African ancestry for whom the utility of PHS46+African may differ. Genotypic data were obtained from PRACTICAL consortium for 6,253 men with African genetic ancestry. Genetic variation in a window spanning 3 African-specific 8q24 SNPs was estimated using 93 PCs. A Cox proportional hazards framework was used to identify the pair of PCs most strongly associated with performance of PHS46+African. A calibration factor (CF) was formulated using estimated Cox coefficients to quantify the extent to which the performance of PHS46+African varies with PC. CF of PHS46+African was strongly associated with the first and twentieth PCs. Predicted CF ranged from 0.41 to 2.94, suggesting that PHS46+African may be up to 7 times more beneficial to some African men than others. The explained relative risk for PHS46+African varied from 3.6% to 9.9% for individuals with low and high CF values, respectively. By cross-referencing our dataset with 1000 Genomes, we identified statistically significant associations between continental and calibration groupings. In conclusion, we identified PCs within 8q24 SNP window that were strongly associated with performance of PHS46+African. Further research to improve clinical utility of polygenic risk scores (or models) is needed to improve health outcomes for men of African ancestry

## Introduction

Polygenic hazard score (PHS) models can test for associations between genetic variants and the age at diagnosis of prostate cancer^1,2^. These models generate personalized estimates of risk that can be used to guide decisions on whether and when to offer screening to men^3–5^. However, development of polygenic models have often included only individuals of European genetic ancestry^6,7^, which could potentially lead to greater prostate cancer disparities. This is a particular concern for prostate cancer. For example, African American men are more likely than other men in the United States to develop prostate cancer, have a younger age of diagnosis, and are more than twice as likely to die from their prostate cancer as white men^8^.

PHS46, a model for prostate cancer trained exclusively in men of European ancestry, was found to be roughly half as effective in African men as in Europeans and Asians^9^. Similar trends were observed for other polygenic scores^6^, highlighting the need for increased diversification of large-scale genome studies in order to address this disparity^7^ in clinical utility. Furthermore, studies have suggested that inequities in the performance of polygenic risk scores may exacerbate disparities for individuals and communities that are already under-represented in research^10^. In an effort to develop more equitable PHS models, our group recently developed an African-ancestry-specific PHS model (PHS46+African)^11^, that substantially improved upon the performance of PHS46.

PHS46+African consists of 46 single nucleotide polymorphisms (SNPs) that were identified in Europeans, together with 3 additional SNPs located on the 8q24 chromosome that were found to uniquely improve performance in men of African genetic ancestry (African men). However, given the inherent genetic diversity on the African continent and gene flow associated with the African diaspora^12^, we believe that PHS46+African may benefit certain subpopulations of African men over others. This would have implications for the general utility of PHS46+African and provide an opportunity to further improve the model.

Therefore, we used principal component (PC) analysis to estimate the genetic relatedness^13,14^ of a dataset of men of African genetic ancestry and to determine whether the performance of PHS46+African varied along PCs representing local genetic ancestry near African-specific SNPs. The PC analysis was conducted on a SNP window encompassing the 3 African-specific SNPs and limited to the 8q24 chromosome. In this way, the African men are projected onto axes, where proximity is based on patterns of genetic variation within this specific section of the genome and agnostic to geographical or social groupings in the dataset. We believe that this “local PC” approach^13^, focusing on 8q24, may uncover subpopulations of African men for whom the utility of PHS46+African differs.

## Material and Methods

### African-Ancestry Dataset

Genotypic and phenotypic data for this study were obtained from the Prostate Cancer Association Group to Investigate Cancer Associated Alterations in the Genome (PRACTICAL)^15^ consortium. Genotyping was performed using the OncoArray^15,16^ chip and covered 444,323 SNPs across the genome. Genotypic data was coded as effect allele counts (0, 1, or 2) for each SNP. Phenotypic data consisted of the prostate cancer case/control status, age at diagnosis, age at last follow-up, genome-wide principal components, and genotypic ancestry.

The genotypic ancestry of each individual was previously estimated using 2,318 ancestry informative markers spanning the entire genome^17^.Ancestral groupings using SNP markers showed good agreement with self-reported race/ethnicity^9^. In total, 6,253 men were classified as having African genotypic ancestry and were used for this analysis. All contributing studies were approved by the relevant ethics committees; written informed consent was obtained from the study participants.

PHS46+African was estimated for each African individual using the 49 SNPs and their respective PHS46+African SNP coefficients from the literature^18^.

### Principal components of 8q24 SNP window

Genetic variation within a narrow window surrounding the three 8q24 SNPs was quantified using principal component decomposition. An initial selection window of SNPs was identified as all SNPs on chromosome 8 lying between the three African-specific 8q24 SNPs in PHS46+African or within 15 kbp of those SNPs in either direction (Figure 1). SNPs were subsequently removed from this window if the call rate was less than 0.95 or if the SNPs could not be cross-referenced against those available on the 1000 Genomes Project Phase 3 dataset^19^. In total, 93 SNPs (Supplementary Table 1) met the selection criteria and constituted the local 8q24 window. Missing SNP calls were replaced with the mean of the genotyped data for that SNP in the African dataset^11,20^. Genetic count data were first standardized across the dataset (mean of 0, standard deviation of 1) before the first 93 principal components (PC) were estimated using the “pca” function in MATLAB.

**Table 1.** Coefficients of Cox model estimating interactions between PC and PHS46+African. Estimates of coefficients from the Cox proportional hazards model describing the association between age of onset of any prostate cancer as the time-to-event, and interactions between 8q24 PCs (PC-1, PC-20) and PHS46+African as the predictors. The model also included interaction terms between the first 4 global ancestry PCs and PHS46+African as covariables.

| Coefficient | Associated predictor | Estimate | p-value |
| --- | --- | --- | --- |
| $\gamma_1$ | PHS46+African | 1.55 | < 1E-16 |
| $\gamma_2$ | PC-1 x PHS46+African | -0.049 | 7.36 E-6 |
| $\gamma_3$ | PC-20 x PHS46+African | 0.21 | 5.84 E-5 |
| $\gamma_4$ | Global Ancestry PC(1-4) x PHS46+African | 7.34 | 0.29 |
| $\gamma_5$ | | -14.71 | 0.33 |
| $\gamma_6$ | | -3.97 | 0.42 |
| $\gamma_7$ | | -2.86 | 0.54 |

**Figure 1.**
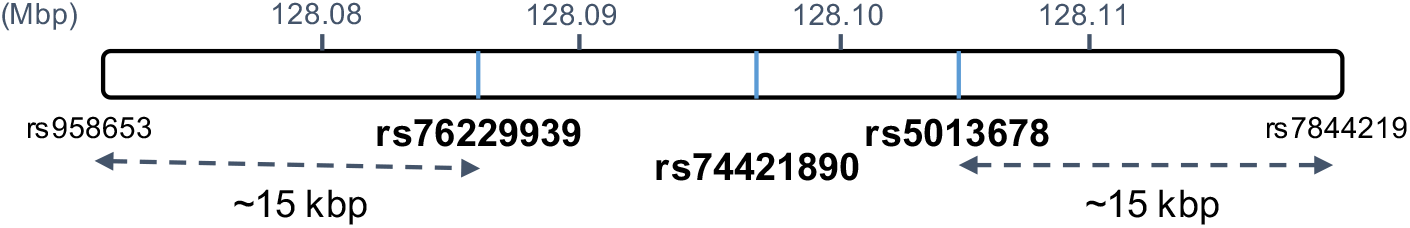
Definition of 8q24 SNP window. A window spanning roughly 15 kbp in either direction from the three 8q24 SNPs (rs76229939, rs74421890, rs5013678) was defined as the 8q24 SNP window.

### Interaction between 8q24 SNP window and PHS46+African

For each principal component (PC) of the 8q24 SNP window, a Cox proportional hazards model was estimated using the age of onset of any prostate cancer as the time-to-event (Eq. 1)

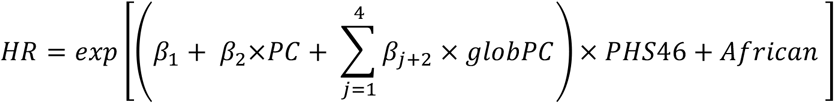

where HR is the hazard rate, β_1_is the coefficient of PHS46+African, β_2_ is the coefficient of the interaction terms between the 8q24 PC and PHS46+African, and β_3_ through β_6_are the coefficients for the interaction terms between the first 4 global ancestry principal components (globPC) and PHS46+African. The model was estimated using the entire African dataset, where each observation was weighted by a sample-weighting correction factor^11,20^ to account for differences between the case-control rate of our dataset and that observed in the general population. Controls were censored at the age of last follow-up. The p-value associated with β_2_ was recorded for each of the 90 PCs from 8q24, after which the two principal components with the smallest p-values 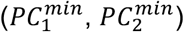 were selected for further analysis. A Cox proportional hazards model was then estimated using both of the selected principal components (Eq. 2)

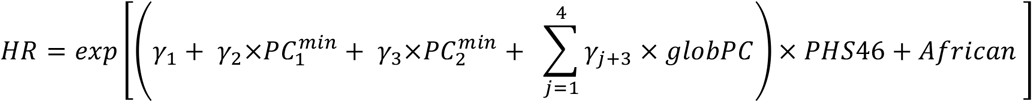

The calibration factor (CF) for PHS46+African, as a function of 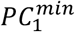 and 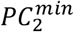, was defined as (Eq. 3)

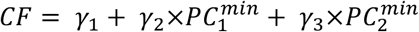

CF was formulated as the performance metric of interest in this study and can be used to quantify the variation in the coefficient of PHS46+African as a result of differences in the expression of variants within the 8q24 SNP window. Larger values of CF suggest stronger associations between the PHS46+African score and the age at diagnosis of prostate cancer. Individuals in the top 33^rd^, middle 33^rd^, and bottom 33^rd^ percentiles of CF values were grouped into high-, middle- and low-calibration groups respectively. These groups were used to facilitate comparisons in CF between PHS models and dataset variables.

### Comparison between PHS46 and PHS46+African in calibration groups

Within each calibration group, a Cox proportional hazards model was fit using the age of diagnosis of any prostate cancer as the time-to-event and either PHS46+African or PHS46 as the sole predictor variable. PHS46 was estimated using SNP coefficients published previously^18^. Controls were censored at age at last follow-up. Models were fit using data only from that calibration group. For each of these group-based models, the explained relative risk (ERR)^21^ was estimated as a measure of model goodness-of-fit. ERR was compared between the two PHS models to determine whether the improvement in performance between the models was evenly distributed across the calibration groups. Empirical confidence intervals for ERR were estimated using 1000 bootstrapped samples.

### Variation in dataset variables across calibration groups

In order to identify differences in characteristics between calibration groups, a set of generalized linear models were fit to study the association between dataset variables (genetic count of three African-specific 8q24 SNPs, case-control status, and age at diagnosis of cases) and calibration groups. In each case, the dataset variable was set as the independent variable, while the calibration group classification (low/middle/high) was set as the predictor. Identity link functions were used for the continuous dataset variables (genetic count of three 8q24 SNPs, age at diagnosis of cases), whereas the logit link function was used for the binary variable (case-control status). Fitted models were then used to predict mean values of the dataset variables for each 2-PC region. In addition, a chi-squared test was used to determine whether any association existed between the contributing study (18 in total) and calibration group.

### Continental differences between calibration groups

The 1000 Genomes (1000G) dataset was used to identify potential differences in continental origins across calibration groups. The 1000G dataset is subdivided into 5 continental groups: European, East Asian, admixed American, South Asian, and African individuals. Genetic counts for the 93-SNP 8q24 window for 2,504 individuals from the 1000G dataset were obtained from publicly available sources^19^. 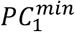and 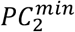scores were estimated using the same scaling factors and PC coefficients derived from the African dataset. CF and calibration groups for each individual in the 1000G dataset were estimated using Equation 3, and the percentile cutoffs derived from the African dataset. Chi-squared tests were used to test for associations between continental and calibration groups.

## Results

### Model interaction between 8q24 PCs and PHS46+African

The first (PC-1) and twentieth (PC-20) principal components had the two smallest p-values (Supplementary Table 2) when estimating the Cox proportional hazards models, as formulated in Eq.1, for all 93 principal components of the 8q24 SNP window. These two principal components were thus selected as 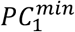and 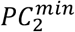and used to estimate the Cox proportional hazards model formulated in Eq. 2. (Table 1). No significant interactions (α = 0.05) were detected between global ancestry principal components and PHS46+African.

**Table 2.** ERR for PHS46+African and PHS46. Mean explained relative risk (ERR) values were tabulated for each of calibration groups using either PHS46 or PHS46+African models. No statistically significant differences in absolute improvement in ERR were observed for any of the calibration groups. Values are tabulated as mean [95% confidence interval].

| Calibration group | PHS46+African | PHS46 | Improvement in ERR |
| --- | --- | --- | --- |
| low | 0.036 [0.019, 0.059] | 0.013 [0.004, 0.028] | 0.023 [0.011, 0.039] |
| middle | 0.056 [0.036, 0.083] | 0.018 [0.007, 0.036] | 0.037 [0.021, 0.055] |
| high | 0.099 [0.068, 0.135] | 0.041 [0.022, 0.069] | 0.058 [0.035, 0.083] |

Principal component coefficients and scaling factors needed to estimate PC-1 and PC-20 from the genetic counts of the 93 SNP-8q24 window are reported in the Supplementary Material (Supplementary Table 3).

### Calibration factor for PHS46+African

CF was plotted as a function PC-1 and PC-20 for every African individual in our dataset (Figure 2A). Qualitatively, the calibration factor tended to increase from high-PC-1/low-PC-20 to low-PC-1/high-PC-20 values from a minimum value of 0.41 to a maximum of 2.94. Grouping the African individuals based on percentiles of CF (Figure 2B) demonstrates this pattern more conspicuously, as the middle calibration group clusters on the PC-1/PC-20 space as a diagonal band that neatly divides the high and low calibration groups.

**Figure 2.**
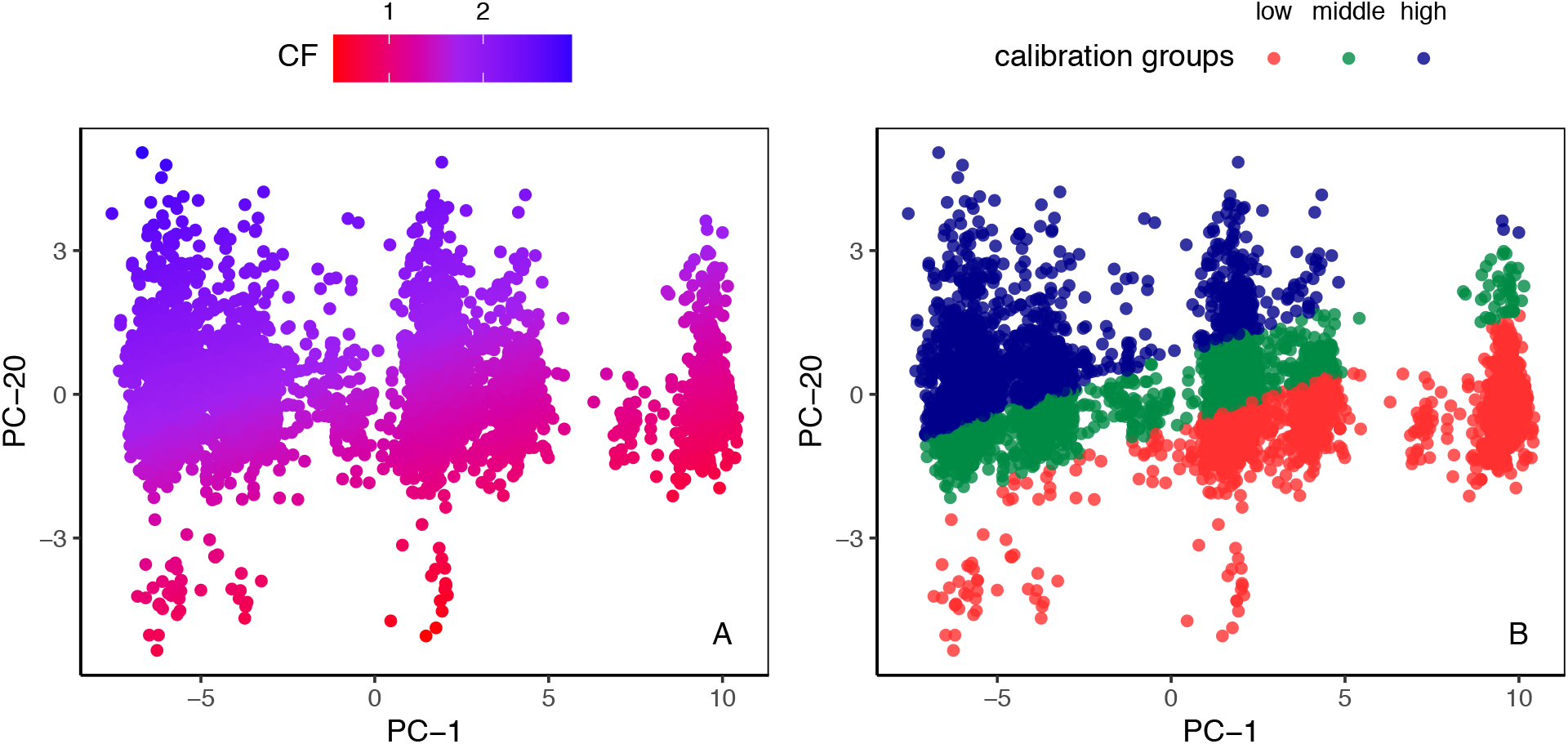
A. CF as a function of PC-1 and PC-20. Calibration factor (CF, Eq. 3) plotted as a function of PC-1 and PC-20. Each point represents an African individual. The calibration factor tended to increase from high-PC-1/low-PC-20 (bottom right corner) to low-PC-1/high-PC-20 (top left corner) values. **B. Calibration groups as a function of PC-1 and PC-20**. Individuals from the African dataset classified into low-(red points, minimum to 33^rd^ percentile), middle-(green points, 33^rd^ to 67^th^ percentile), and high-(blue points, 67^th^ to maximum) calibration groups.

### Comparing explained relative risk (ERR) between calibration groups and across PHS models

Mean ERR values for both PHS46 and PHS46+African were greater for the high calibration group than for the low calibration group (Table 2). Improvement of ERR with addition of the three African-specific SNPs was estimated as the as the difference between the ERR values of PHS46+African and PHS46. The absolute improvement in ERR was comparable for all three calibration groups.

### Variation in dataset variables across 2-PC space

Fitted generalized linear models were used to predict mean values of case-fraction, age of cases, and genetic counts of three 8q24 SNPs for the calibration groups (Supplementary Table 4). No significant (p < 0.05) differences were found in the predicted age of cases across the groups. The predicted fraction of cases in the high-calibration group was lower (0.49) than that of the low-calibration group (0.53). The predicted mean genetic count of rs5013678 in the high-calibration group (0.27) was greater than that in the low-calibration (0.092) and middle-calibration (0.17) groups. No statistically significant association was detected between contributing study and calibration group (Supplementary Table 5).

As a post-hoc analysis, R^2^ values were estimated between case-control status and CF (0.0025) as well as between mean genetic count of rs5013678 and CF (0.039), suggesting that less than 4% of the variation in CF could be explained by each of these variables.

### Continental characterization of calibration groups

The 1000G dataset was mapped into the 2-dimensional space defined by PC-1 and PC-20 and stratified by continental group (Figure 3). In general, individuals from the 1000G dataset mapped within the boundaries defined by the OncoArray African dataset. A statistically significant association (χ^2^ = 288, p < 0.001) was detected between continental and calibration groups in the 1000G dataset. Analysis of the standardized residuals of the test (Supplementary Table 6) revealed that the largest deviation from the expected cross-tabulation counts of the two variables was the greater-than-expected number of African individuals in the low-calibration group. Further analysis of the cross-tabulation between the 2 variables (Supplementary Table 7) revealed that the individuals of European origin made up the largest continental group within the high-calibration individuals (20%).

**Figure 3.**
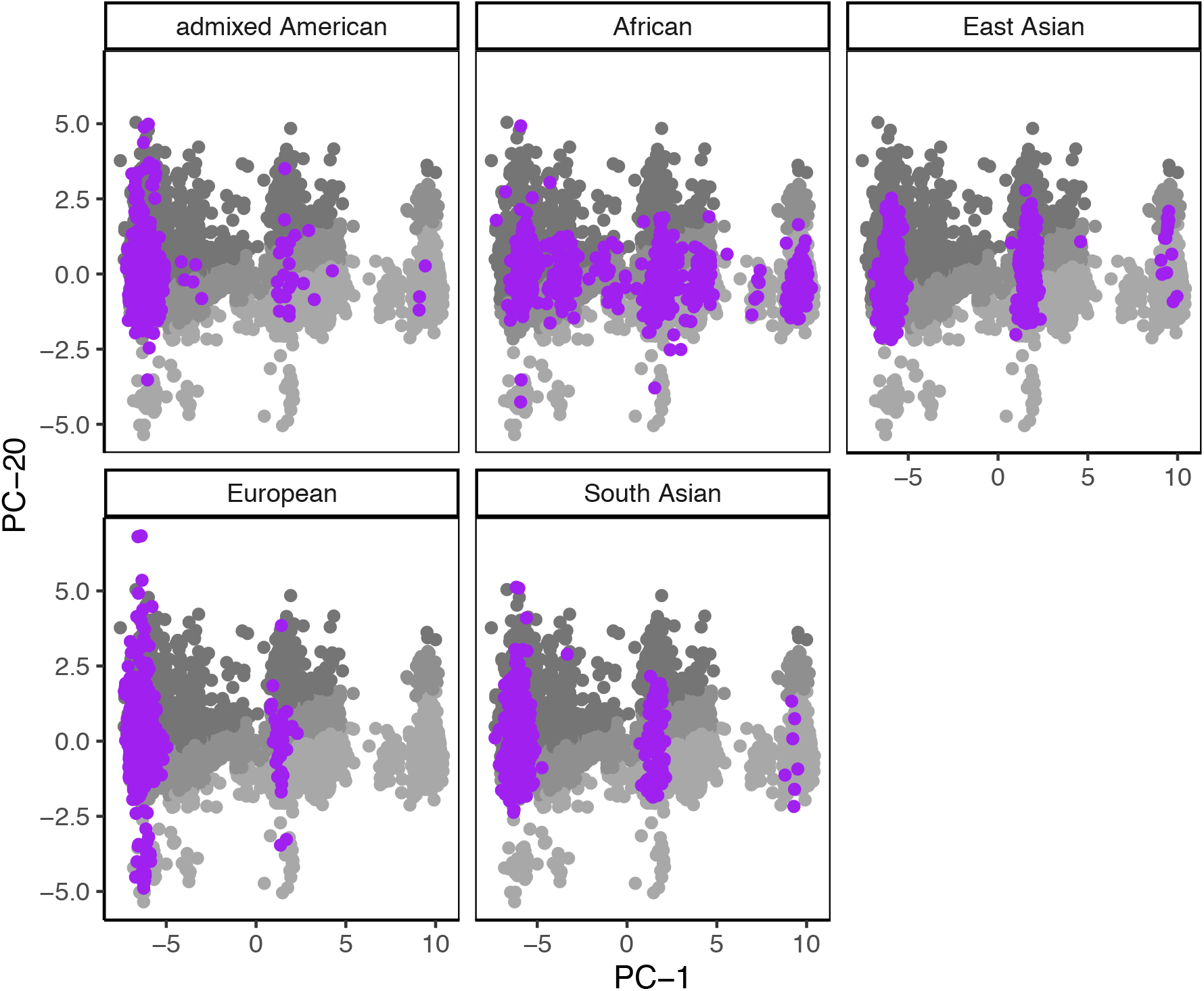
1000G dataset mapped to PC-1 and PC-20. 2,504 individuals from the 1000G dataset (purple dots) were mapped onto the 2-dimensional space defined by PC-1 and PC-20. Each pane represents a different continental group: African, admixed American, East Asian, European, and South Asian. The mapping is overlaid on a grayscale version of Figure 2B (i.e., gray dots represent individuals in the African-Ancestry Dataset used in the present study).

## Discussion

Genetic models developed predominantly with European data may widen health disparities. PHS46+African incorporates African-specific SNPs from 8q24 and improves performance in men of African ancestry, but we have demonstrated here that heterogeneity in local ancestry in 8q24 can affect performance gains.

We identified two principal components within the 8q24 region (PC-1 and PC-20) that were most strongly associated with the calibration factor of PHS46+African among men of African ancestry. The model-predicted CF values ranged from 0.41 to 2.94 for the combinations of PC-1 and PC-20 found in our dataset, suggesting that PHS46+African was roughly 7 times more useful in stratifying risk in some individuals compared to others. Assigning individuals into three equally sized subsets based on thresholds of CF, we identified low-, middle-, and high-calibration groups within our dataset. The goodness-of-fit for PHS46+African in each of these groups, assessed using the explained relative risk, was also found to increase from low-to high-calibration group. Improvements in ERR were observed across all groups when PHS46+African was used instead of PHS46.

The discovery of PC-1 and PC-20 allows us to identify individuals for whom risk stratification using PHS46+African is expected to be less precise. Several strategies could be implemented to improve equity in performance of PHS46+African in men of African genetic ancestry. These strategies can be pursued simultaneously with essential work to increase the overall diversity of genetic studies. For example, efforts to discover additional SNPs in future datasets could include weighting of individuals according to their PC-1 and PC-20 values, in order to enrich discovery of SNPs that preferentially benefit those in the low-calibration group. In addition, SNP weights in PHS46+African might be re-estimated to account for variations in effect sizes with calibration group. The principal components can also be used prospectively to guide enrollment in genome wide association studies so as to ensure that individuals from low-calibration groups are adequately represented in the datasets.

By cross-referencing our dataset with individuals from 1000 Genomes, we identified continent-level differences in calibration groups that suggest ancestral origins for the genetic relatedness defined by PC-1 and PC-20. The mapping of 1000G data into the 2-dimensional space defined by PC-1 and PC-20 may provide clues as to the underlying variation in CF. 46 of the 49 SNPs used in PHS46+African were discovered in a dataset consisting entirely of men with European genetic ancestry. Therefore, it is unsurprising that the high-calibration group overlapped substantially with men of European ancestry, as defined by 1000G. However, certain continental groups that were not explicitly used in training of model weights, such as South Asian and admixed American, also exhibited overlap with the high-calibration group (Supplementary Table 7). On the opposite end of the spectrum, the low-calibration group overlapped with primarily African and East Asian men. Further investigation into pockets of reference groups, from 1000G and other datasets, that share common values of PC-1 and PC-20 may reveal ancestral linkages that may help to predict which ancestral groups or datasets a model is most likely to perform well in.

There are several limitations to the current analysis to consider. We studied the same dataset that was used to identify the 3 African-specific SNPs. Therefore, the dependence of PHS46+African on PC-1 and PC-20 will need to be validated in independent test sets to ensure generalizability of findings. Our study is further limited by a relatively small number of observations compared to those found in larger, often predominantly European, genome-wide association studies. While a larger dataset would allow us to more robustly estimate model coefficients, the mapping of 1000G data into the same 2-dimensional space suggests that our current sample accurately portrays the extent of variation in the 8q24 SNP window (Figure 3). In addition, the calibration groups were selected based on arbitrary thresholds of CF and were used solely to simplify the analysis. Further investigations will be required to determine whether distinct subpopulations, based on ancestral relatedness, may exist in the 2-dimensional PC space.

In conclusion, we used local PC analysis to identify axes of variation within the 8q24 SNP window that were strongly associated with the performance of PHS46+African. Mapping our dataset onto these axes revealed that PHS46+African may meaningfully underperform in certain individuals of African genetic ancestry. Investigation into the origins of both high- and low-performing groups can be used to generate a model that is more equitable in performance across subpopulations of men with African ancestry.

### Ethics Statement

The present analyses used de-identified data from the PRACTICAL consortium and have been approved by the Institutional Review Board at the University of California San Diego.

## Data Availability

The data used in this work were obtained from the Prostate Cancer Association Group to Investigate Cancer Associated Alterations in the Genome (PRACTICAL) consortium. 
Readers who are interested in accessing the data must first submit a proposal to the Data Access Committee. If the reader is not a member of the consortium, their concept form must be sponsored by a principal investigator (PI) of one of the PRACTICAL consortium member studies. If approved by the Data Access Committee, PIs within the consortium, each of whom retains ownership of their data submitted to the consortium, can then choose to participate in the specific proposal.  In addition, portions of the data are available for request from dbGaP (database of Genotypes and Phenotypes) which is maintained by the National Center for Biotechnology Information (NCBI): https://www.ncbi.nlm.nih.gov/gap/?term=Icogs+prostatehttps://www.ncbi.nlm.nih.gov/gap/?term=Icogs+prostate. 
Anyone can apply to join the consortium. The eligibility requirements are listed here: http://practical.icr.ac.uk/blog/?page_id=9. Joining the consortium would not guarantee access, as a proposal for access would still be submitted to the Data Access Committee, but there would be no need for a separate member sponsor. Readers may find information about application by using the contact information below:
Rosalind Eeles
Principal Investigator for PRACTICAL
Professor of Oncogenetics
Institute of Cancer Research (ICR)
Sutton, UK
URL: http://practical.icr.ac.uk
Tel: ++44 (0)20 8722 4094

## Conflict of Interest

All authors declare no support from any organization for the submitted work except as follows: AMD and TMS report a research grant from the US Department of Defense. OAA reports research grants from KG Jebsen Stiftelsen, Research Council of Norway, and South East Norway Health Authority.

Authors declare no financial relationships with any organizations that might have an interest in the submitted work in the previous three years except as follows, with all of these relationships outside the present study: TMS reports honoraria from Multimodal Imaging Services Corporation for imaging segmentation, honoraria from WebMD, Inc. for educational content, as well as a past research grant from Varian Medical Systems. OAA reports speaker honoraria from Lundbeck.

Authors declare no other relationships or activities that could appear to have influenced the submitted work except as follows: OAA has a patent application # U.S. 20150356243 pending; AMD also applied for this patent application and assigned it to UC San Diego. AMD has additional disclosures outside the present work: founder, equity holder, and advisory board member for CorTechs Labs, Inc.; founder and equity holder in HealthLytix, Inc., advisory board member of Human Longevity, Inc.; recipient of nonfinancial research support from General Electric Healthcare. OAA is a consultant for HealthLytix, Inc.

Additional acknowledgments for the PRACTICAL consortium and contributing studies are described in the Appendix A3

## Funding

This study was funded in part by grants from the University of California (#C21CR2060), the United States National Institute of Health/National Institute of Biomedical Imaging and Bioengineering (#K08EB026503), the Research Council of Norway (#223273), KG Jebsen Stiftelsen, and South East Norway Health Authority.

Funding for the PRACTICAL consortium member studies is detailed in the Appendix A2.

The content is solely the responsibility of the authors and does not necessarily represent the official views of any of the funding agencies, who had no role in the design and conduct of the study; collection, management, analysis, and interpretation of the data; preparation, review, or approval of the manuscript; and decision to submit the manuscript for publication.

## Data Availability Statement

The data used in this work were obtained from the Prostate Cancer Association Group to Investigate Cancer Associated Alterations in the Genome (PRACTICAL) consortium.

Readers who are interested in accessing the data must first submit a proposal to the Data Access Committee. If the reader is not a member of the consortium, their concept form must be sponsored by a principal investigator (PI) of one of the PRACTICAL consortium member studies. If approved by the Data Access Committee, PIs within the consortium, each of whom retains ownership of their data submitted to the consortium, can then choose to participate in the specific proposal. In addition, portions of the data are available for request from dbGaP (database of Genotypes and Phenotypes) which is maintained by the National Center for Biotechnology Information (NCBI): https://www.ncbi.nlm.nih.gov/gap/?term=Icogs+prostatehttps://www.ncbi.nlm.nih.gov/gap/?term=Icogs+prostate.

Anyone can apply to join the consortium. The eligibility requirements are listed here: http://practical.icr.ac.uk/blog/?page_id=9. Joining the consortium would not guarantee access, as a proposal for access would still be submitted to the Data Access Committee, but there would be no need for a separate member sponsor. Readers may find information about application by using the contact information below

Rosalind Eeles

Principal Investigator for PRACTICAL Professor of Oncogenetics

Institute of Cancer Research (ICR) Sutton, UK

URL http://practical.icr.ac.uk Tel: ++44 (0)20 8722 4094

