## Supplementary Data for "Performance of African-ancestry-specific polygenic hazard score varies according to local ancestry in 8q24"

**Table of Contents**


| | |
|---|---|
| Supplemental Table 1 Local neighborhood SNP window | 3 |
| Supplementary Table 2. Interaction term between 8q24 PC and PHS46+African | 6 |
| Supplementary Table 3. Estimating PC-1 and PC-20 | 9 |
| Supplementary Table 4. Predicted dataset variables by calibration group | 12 |
| Supplementary Table 5. Contributing study by calibration group | 13 |
| Supplementary Table 6. Standardized chi-squared residuals | 14 |
| Supplementary Table 7. Cross-tabulation of continental and calibration groups | 15 |
| Appendix 1. Members of the PRACTICAL Consortium | 16 |
| Appendix 2. Funding sources for the PRACTICAL consortium | 21 |

**Supplemental Table 1 Local neighborhood SNP window.** RS number, chromosome, position (version 37), effect allele, reference allele and effect allele frequency in Africans from the 1000G dataset for the 93 SNPs that constitute 8q24 SNP window.

| RS number | Chromosome | Position | Effect | Reference | Effect Allele Frequency |
|---:|---:|---:|---:|---:|---:|
| rs958653 | 8 | 128070669 | G | A | 0.072 |
| rs9297751 | 8 | 128070847 | C | T | 0.386 |
| rs34241583 | 8 | 128070900 | A | G | 0.209 |
| rs10505485 | 8 | 128071064 | G | A | 0.272 |
| rs9649957 | 8 | 128071305 | T | C | 0.290 |
| rs58120361 | 8 | 128071380 | T | C | 0.434 |
| rs62529863 | 8 | 128071464 | T | C | 0.385 |
| rs17763940 | 8 | 128071504 | A | G | 0.012 |
| rs6983515 | 8 | 128071922 | A | G | 0.467 |
| rs7833560 | 8 | 128072019 | C | T | 0.467 |
| rs7003994 | 8 | 128072064 | C | T | 0.023 |
| rs1456314 | 8 | 128075690 | A | G | 0.530 |
| rs16901935 | 8 | 128076545 | G | A | 0.047 |
| rs77541621 | 8 | 128077146 | A | G | 0.000 |
| rs12542102 | 8 | 128077521 | C | T | 0.653 |
| rs2392729 | 8 | 128079666 | T | C | 0.039 |
| rs77120121 | 8 | 128079697 | T | C | 0.036 |
| rs7824674 | 8 | 128081093 | G | A | 0.097 |
| rs7824923 | 8 | 128081138 | T | C | 0.097 |
| rs7825394 | 8 | 128081449 | T | C | 0.097 |
| rs61386925 | 8 | 128083486 | G | A | 0.058 |
| rs6993569 | 8 | 128084097 | A | G | 0.489 |
| rs6994316 | 8 | 128084539 | G | A | 0.206 |
| rs76229939 | 8 | 128085394 | G | A | 0.048 |
| rs114798100 | 8 | 128085434 | G | A | 0.048 |
| rs11998248 | 8 | 128085755 | A | G | 0.483 |
| rs35450468 | 8 | 128086032 | G | A | 0.489 |
| rs74634260 | 8 | 128086189 | C | A | 0.002 |
| rs76595456 | 8 | 128087829 | T | C | 0.115 |
| rs6470494 | 8 | 128087904 | T | C | 0.292 |
| rs7843737 | 8 | 128088066 | A | G | 0.293 |
| rs75500912 | 8 | 128089536 | C | A | 0.081 |
| rs4581008 | 8 | 128090865 | T | C | 0.281 |
| rs36002700 | 8 | 128091158 | G | T | 0.157 |
| rs12544839 | 8 | 128091382 | A | G | 0.328 |
| rs72725868 | 8 | 128091418 | G | A | 0.004 |

| | | | | | |
|---|---|---|---|---|---|
| rs10956351 | 8 | 128091559 | G | A | 0.581 |
| rs1016342 | 8 | 128092455 | T | C | 0.676 |
| rs73705708 | 8 | 128092911 | G | A | 0.141 |
| rs1031589 | 8 | 128092982 | G | A | 0.284 |
| rs1031588 | 8 | 128093277 | A | C | 0.026 |
| rs1016343 | 8 | 128093297 | T | C | 0.210 |
| rs4871008 | 8 | 128093541 | T | C | 0.284 |
| rs7001504 | 8 | 128094483 | T | G | 0.284 |
| rs13252298 | 8 | 128095156 | G | A | 0.030 |
| rs73705712 | 8 | 128095977 | G | A | 0.117 |
| rs74421890 | 8 | 128096183 | T | C | 0.042 |
| rs59765225 | 8 | 128099765 | G | A | 0.168 |
| rs9656814 | 8 | 128100977 | T | C | 0.125 |
| rs11994653 | 8 | 128101343 | G | A | 0.656 |
| rs7000321 | 8 | 128103019 | C | A | 0.214 |
| rs1456315 | 8 | 128103937 | T | C | 0.581 |
| rs5013678 | 8 | 128103979 | C | T | 0.081 |
| rs118005113 | 8 | 128104745 | A | C | 0.001 |
| rs1073997 | 8 | 128105187 | C | A | 0.470 |
| rs57775023 | 8 | 128105672 | A | G | 0.077 |
| rs7016830 | 8 | 128106235 | G | A | 0.465 |
| rs115266272 | 8 | 128106397 | A | G | 0.033 |
| rs6983561 | 8 | 128106880 | C | A | 0.526 |
| rs16901949 | 8 | 128107153 | C | A | 0.371 |
| rs60681470 | 8 | 128107192 | A | G | 0.504 |
| rs16901950 | 8 | 128107243 | A | G | 0.370 |
| rs16901952 | 8 | 128107270 | C | T | 0.370 |
| rs16901953 | 8 | 128108229 | C | T | 0.381 |
| rs146278672 | 8 | 128108324 | T | C | 0.033 |
| rs35365584 | 8 | 128108726 | A | G | 0.098 |
| rs17832285 | 8 | 128108993 | G | A | 0.092 |
| rs7825340 | 8 | 128109129 | G | A | 0.091 |
| rs12544977 | 8 | 128109287 | A | G | 0.198 |
| rs73344342 | 8 | 128109356 | A | G | 0.380 |
| rs16901959 | 8 | 128109530 | G | A | 0.381 |
| rs7826337 | 8 | 128109574 | A | G | 0.616 |
| rs7826388 | 8 | 128109776 | T | C | 0.209 |
| rs7830341 | 8 | 128109930 | A | G | 0.380 |
| rs16901966 | 8 | 128110252 | G | A | 0.381 |
| rs16901967 | 8 | 128110277 | G | A | 0.379 |
| rs7001069 | 8 | 128110646 | G | A | 0.380 |
| rs11781162 | 8 | 128110896 | G | A | 0.001 |
| rs73348236 | 8 | 128111470 | T | C | 0.380 |

| | | | | | |
|---|---|---|---|---|---|
| rs57010632 | 8 | 128111717 | G | A | 0.531 |
| rs16901969 | 8 | 128112097 | C | A | 0.380 |
| rs61265543 | 8 | 128112446 | T | C | 0.531 |
| rs16901970 | 8 | 128112715 | G | T | 0.381 |
| rs10453084 | 8 | 128112779 | A | G | 0.380 |
| rs6987723 | 8 | 128112859 | A | G | 0.380 |
| rs113694160 | 8 | 128112976 | A | G | 0.002 |
| rs7824785 | 8 | 128114710 | T | C | 0.379 |
| rs73348257 | 8 | 128115259 | A | G | 0.379 |
| rs75170735 | 8 | 128115378 | G | A | 0.001 |
| rs28686871 | 8 | 128116480 | T | G | 0.032 |
| rs1456306 | 8 | 128116500 | A | G | 0.106 |
| rs57462547 | 8 | 128117863 | T | C | 0.535 |
| rs7844219 | 8 | 128118815 | G | A | 0.536 |

**Supplementary Table 2. Interaction term between 8q24 PC and PHS46+African.** P-values of the coefficient associated with the interaction term between 8q24 principal components and PHS46+African ($\beta_2$ from Equation 1) are tabulated in ascending order. PC-1 and PC-20 (highlighted in gray) were associated with the two smallest p-values and therefore, selected for further analysis.

| Principal component | p-value of $\beta_2$ – PC x PHS46+African |
|---|---|
| 1 | 1.06E-05 |
| 20 | 8.52E-05 |
| 90 | 1.34E-04 |
| 84 | 2.77E-04 |
| 56 | 5.64E-04 |
| 51 | 7.03E-04 |
| 93 | 1.63E-03 |
| 82 | 2.55E-03 |
| 78 | 6.06E-03 |
| 73 | 6.60E-03 |
| 31 | 9.85E-03 |
| 57 | 1.44E-02 |
| 11 | 1.82E-02 |
| 12 | 1.92E-02 |
| 71 | 2.65E-02 |
| 68 | 2.65E-02 |
| 38 | 3.51E-02 |
| 74 | 3.73E-02 |
| 8 | 3.75E-02 |
| 92 | 3.84E-02 |
| 65 | 3.98E-02 |
| 23 | 4.01E-02 |
| 14 | 4.14E-02 |
| 62 | 4.15E-02 |
| 88 | 4.73E-02 |
| 42 | 6.83E-02 |
| 69 | 7.04E-02 |
| 87 | 7.18E-02 |
| 53 | 7.70E-02 |
| 75 | 7.76E-02 |
| 85 | 8.41E-02 |
| 22 | 1.02E-01 |
| 79 | 1.19E-01 |

| | |
|---|---|
| 81 | 1.31E-01 |
| 2 | 1.32E-01 |
| 35 | 1.78E-01 |
| 24 | 1.85E-01 |
| 58 | 1.85E-01 |
| 83 | 2.09E-01 |
| 32 | 2.18E-01 |
| 9 | 2.28E-01 |
| 55 | 2.33E-01 |
| 41 | 2.55E-01 |
| 91 | 3.15E-01 |
| 13 | 3.18E-01 |
| 59 | 3.40E-01 |
| 66 | 3.73E-01 |
| 40 | 4.13E-01 |
| 72 | 4.23E-01 |
| 43 | 4.27E-01 |
| 18 | 4.29E-01 |
| 19 | 4.31E-01 |
| 63 | 4.32E-01 |
| 37 | 4.32E-01 |
| 3 | 4.42E-01 |
| 76 | 4.46E-01 |
| 6 | 4.62E-01 |
| 50 | 4.69E-01 |
| 60 | 4.94E-01 |
| 33 | 4.95E-01 |
| 70 | 5.13E-01 |
| 54 | 5.13E-01 |
| 36 | 5.17E-01 |
| 52 | 5.20E-01 |
| 27 | 5.37E-01 |
| 10 | 5.50E-01 |
| 16 | 5.53E-01 |
| 44 | 5.55E-01 |
| 61 | 5.69E-01 |
| 86 | 5.70E-01 |
| 77 | 5.97E-01 |
| 28 | 6.27E-01 |
| 29 | 6.39E-01 |

| | |
|---|---|
| 4 | 6.40E-01 |
| 39 | 6.45E-01 |
| 17 | 6.77E-01 |
| 30 | 6.85E-01 |
| 21 | 6.91E-01 |
| 26 | 6.92E-01 |
| 64 | 7.18E-01 |
| 25 | 7.31E-01 |
| 7 | 7.39E-01 |
| 80 | 7.76E-01 |
| 89 | 7.83E-01 |
| 48 | 7.86E-01 |
| 15 | 7.91E-01 |
| 5 | 8.69E-01 |
| 34 | 8.79E-01 |
| 46 | 8.80E-01 |
| 67 | 9.37E-01 |
| 47 | 9.47E-01 |
| 45 | 9.55E-01 |
| 49 | 9.66E-01 |

**Supplementary Table 3. Estimating PC-1 and PC-20.** Scaling factors and PC coefficients used to estimate PC-1 and PC-20 from the genetic counts of the 93-SNP 8q24 window.

| RS number | Scaling factor | | Coefficients | |
|---|---|---|---|---|
| | Mean | Std Dev | PC-1 | PC-20 |
| rs958653 | 0.205 | 0.430 | -0.026 | -0.162 |
| rs9297751 | 0.732 | 0.684 | -0.015 | -0.087 |
| rs34241583 | 0.460 | 0.590 | -0.007 | 0.057 |
| rs10505485 | 0.524 | 0.615 | 0.017 | 0.220 |
| rs9649957 | 0.647 | 0.657 | 0.001 | 0.156 |
| rs58120361 | 0.849 | 0.709 | -0.002 | 0.036 |
| rs62529863 | 0.758 | 0.688 | -0.016 | 0.061 |
| rs17763940 | 0.058 | 0.240 | -0.031 | 0.043 |
| rs6983515 | 0.849 | 0.694 | -0.030 | -0.136 |
| rs7833560 | 0.856 | 0.704 | -0.031 | -0.139 |
| rs7003994 | 0.050 | 0.224 | 0.041 | -0.092 |
| rs1456314 | 1.045 | 0.714 | 0.028 | -0.067 |
| rs16901935 | 0.087 | 0.289 | -0.015 | 0.348 |
| rs77541621 | 0.009 | 0.094 | 0.009 | 0.046 |
| rs12542102 | 1.223 | 0.697 | 0.037 | -0.068 |
| rs2392729 | 0.180 | 0.410 | -0.052 | -0.027 |
| rs77120121 | 0.092 | 0.299 | 0.013 | -0.073 |
| rs7824674 | 0.294 | 0.500 | -0.055 | -0.092 |
| rs7824923 | 0.293 | 0.499 | -0.055 | -0.093 |
| rs7825394 | 0.293 | 0.499 | -0.055 | -0.093 |
| rs61386925 | 0.154 | 0.379 | -0.049 | -0.223 |
| rs6993569 | 0.923 | 0.715 | 0.004 | 0.019 |
| rs6994316 | 0.411 | 0.575 | 0.019 | -0.048 |
| rs76229939 | 0.125 | 0.341 | 0.002 | 0.011 |
| rs114798100 | 0.125 | 0.341 | 0.002 | 0.011 |
| rs11998248 | 0.900 | 0.714 | -0.001 | 0.011 |
| rs35450468 | 0.922 | 0.715 | 0.004 | 0.018 |
| rs74634260 | 0.006 | 0.079 | -0.008 | 0.151 |
| rs76595456 | 0.248 | 0.463 | 0.006 | -0.060 |
| rs6470494 | 0.599 | 0.648 | 0.020 | 0.033 |
| rs7843737 | 0.583 | 0.642 | 0.013 | 0.038 |
| rs75500912 | 0.182 | 0.406 | 0.015 | -0.102 |
| rs4581008 | 0.581 | 0.633 | 0.012 | 0.038 |
| rs36002700 | 0.266 | 0.481 | -0.034 | -0.055 |
| rs12544839 | 0.647 | 0.647 | 0.027 | 0.041 |
| rs72725868 | 0.024 | 0.156 | -0.021 | 0.157 |
| rs10956351 | 1.104 | 0.694 | 0.005 | -0.039 |
| rs1016342 | 1.293 | 0.675 | -0.016 | -0.040 |

| | | | | |
|---|---|---|---|---|
| rs73705708 | 0.293 | 0.499 | -0.001 | -0.048 |
| rs1031589 | 0.597 | 0.649 | 0.000 | -0.025 |
| rs1031588 | 0.132 | 0.356 | -0.055 | -0.010 |
| rs1016343 | 0.431 | 0.589 | 0.030 | 0.063 |
| rs4871008 | 0.597 | 0.649 | 0.000 | -0.023 |
| rs7001504 | 0.595 | 0.648 | 0.000 | -0.025 |
| rs13252298 | 0.123 | 0.348 | -0.038 | -0.097 |
| rs73705712 | 0.196 | 0.422 | 0.012 | -0.080 |
| rs74421890 | 0.077 | 0.271 | -0.016 | -0.064 |
| rs59765225 | 0.327 | 0.527 | 0.011 | -0.011 |
| rs9656814 | 0.334 | 0.530 | -0.026 | -0.039 |
| rs11994653 | 1.204 | 0.696 | -0.002 | -0.001 |
| rs7000321 | 0.457 | 0.591 | 0.032 | 0.072 |
| rs1456315 | 1.150 | 0.696 | 0.090 | 0.064 |
| rs5013678 | 0.178 | 0.396 | -0.048 | 0.028 |
| rs118005113 | 0.005 | 0.071 | -0.008 | -0.026 |
| rs1073997 | 0.831 | 0.700 | 0.180 | 0.005 |
| rs57775023 | 0.211 | 0.434 | -0.060 | 0.038 |
| rs7016830 | 0.824 | 0.700 | 0.182 | -0.006 |
| rs115266272 | 0.103 | 0.311 | -0.011 | 0.047 |
| rs6983561 | 0.988 | 0.705 | 0.167 | 0.026 |
| rs16901949 | 0.686 | 0.657 | 0.192 | 0.008 |
| rs60681470 | 0.931 | 0.708 | 0.174 | 0.009 |
| rs16901950 | 0.665 | 0.666 | 0.193 | -0.001 |
| rs16901952 | 0.670 | 0.667 | 0.194 | -0.002 |
| rs16901953 | 0.685 | 0.671 | 0.196 | -0.015 |
| rs146278672 | 0.103 | 0.311 | -0.011 | 0.046 |
| rs35365584 | 0.240 | 0.460 | -0.068 | -0.016 |
| rs17832285 | 0.194 | 0.418 | -0.060 | 0.012 |
| rs7825340 | 0.206 | 0.429 | -0.059 | 0.054 |
| rs12544977 | 0.406 | 0.570 | -0.085 | -0.019 |
| rs73344342 | 0.681 | 0.670 | 0.195 | -0.015 |
| rs16901959 | 0.685 | 0.671 | 0.196 | -0.016 |
| rs7826337 | 1.179 | 0.693 | 0.138 | -0.010 |
| rs7826388 | 0.393 | 0.564 | 0.138 | 0.012 |
| rs7830341 | 0.680 | 0.670 | 0.195 | -0.016 |
| rs16901966 | 0.683 | 0.671 | 0.196 | -0.017 |
| rs16901967 | 0.683 | 0.671 | 0.196 | -0.017 |
| rs7001069 | 0.683 | 0.671 | 0.196 | -0.017 |
| rs11781162 | 0.007 | 0.082 | -0.010 | 0.265 |
| rs73348236 | 0.683 | 0.671 | 0.196 | -0.017 |
| rs57010632 | 0.997 | 0.703 | 0.170 | 0.007 |
| rs16901969 | 0.676 | 0.672 | 0.195 | -0.018 |

| | | | | |
|---|---|---|---|---|
| rs61265543 | 0.987 | 0.711 | 0.172 | -0.005 |
| rs16901970 | 0.683 | 0.671 | 0.196 | -0.016 |
| rs10453084 | 0.683 | 0.671 | 0.196 | -0.017 |
| rs6987723 | 0.680 | 0.670 | 0.195 | -0.016 |
| rs113694160 | 0.009 | 0.094 | -0.013 | -0.378 |
| rs7824785 | 0.682 | 0.671 | 0.196 | -0.017 |
| rs73348257 | 0.683 | 0.670 | 0.196 | -0.016 |
| rs75170735 | 0.101 | 0.434 | 0.001 | 0.528 |
| rs28686871 | 0.098 | 0.307 | -0.042 | 0.011 |
| rs1456306 | 0.238 | 0.459 | -0.066 | -0.017 |
| rs57462547 | 0.988 | 0.711 | 0.170 | -0.004 |
| rs7844219 | 0.989 | 0.711 | 0.170 | -0.003 |

**Supplementary Table 4. Predicted dataset variables by calibration group.** Predicted mean value of dataset variables (case-rate, age of cases, and genetic count of 8q24 SNPs) for each calibration group using generalized linear models. Table values marked with asterisks are significantly different (p<0.05) than the corresponding value for the low-performing 2-PC region.

| Calibration group | Case-fraction | Age of Cases | Genetic count | | |
|---|---|---|---|---|---|
| | | | rs76229939 | rs74421890 | rs5013678 |
| low | 0.54 | 62.3 | 0.13 | 0.069 | 0.092 |
| medium | 0.53 | 62.3 | 0.13 | 0.080 | 0.17* |
| high | 0.49* | 62.7 | 0.12 | 0.081 | 0.27* |

**Supplementary Table 5. Contributing study by calibration group.** Cross-tabulation of contributing study (18 in total) and calibration group. No statistically significant association (p=0.05) between the two variables was found.

| Calibration group | Contributing Study | | | | |
|---|---|---|---|---|---|
| | BioVU | CeRePP | CPDR | EPICAP | KARUPROSTATE |
| low | 78 | 44 | 62 | 8 | 240 |
| medium | 64 | 61 | 61 | 13 | 276 |
| high | 62 | 80 | 53 | 8 | 233 |
| | MIAMI-WFPCS | MOFFITT | NMHS | PCaP | PROtEuS |
| low | 28 | 57 | 136 | 349 | 41 |
| medium | 37 | 61 | 119 | 332 | 36 |
| high | 43 | 74 | 109 | 286 | 46 |
| | SABOR | SCCS | SCPCS | SFPCS | SWOG-PCPT |
| low | 70 | 568 | 26 | 38 | 57 |
| medium | 78 | 586 | 30 | 46 | 59 |
| high | 63 | 635 | 33 | 33 | 48 |
| | SWOG-SELECT | UKGPCS | WUGS | | |
| low | 70 | 127 | 64 | | |
| medium | 66 | 123 | 79 | | |
| high | 61 | 115 | 81 | | |

**Supplementary Table 6. Standardized chi-squared residuals.** Standardized residuals of the chi-squared test between continental and calibration groups in the 1000 Genomes dataset are tabulated. The largest contribution to the statistically significant chi-squared value was a result of the greater-than-expected number of Africans within the low-calibration group.

| | | Calibration group | | |
|---|---|---|---|---|
| | | low | middle | high |
| Continental group | African | 14.3 | -8.8 | -8.7 |
| | American | -7.9 | 5.6 | 3.7 |
| | Asian | 2.7 | -1.2 | -2.4 |
| | European | -7.7 | 4.3 | 5.4 |
| | South Asian | -4.0 | 1.8 | 3.3 |

**Supplementary Table 7. Cross-tabulation of continental and calibration groups.** Cross-tabulation of continental and calibration groups by calibration group (column sum equals 1), and by continental group (row sum equals 1). Values are expressed as percentages.

| By calibration group | | Calibration group | | |
|---|---|---|---|---|
| | | low | middle | high |
| Continental group | African | 38.8 | 16.1 | 6.5 |
| | American | 8.5 | 19.0 | 20.5 |
| | Asian | 22.3 | 18.9 | 15.2 |
| | European | 14.0 | 24.6 | 31.4 |
| | South Asian | 16.4 | 21.4 | 26.4 |

| By continental group | | Calibration group | | |
|---|---|---|---|---|
| | | low | middle | high |
| Continental group | African | 74.6 | 22.2 | 3.2 |
| | American | 31.1 | 49.9 | 19.0 |
| | Asian | 56.2 | 34.1 | 9.7 |
| | European | 35.4 | 44.5 | 20.1 |
| | South Asian | 42.7 | 39.9 | 17.4 |

### Appendix 1. Members of the PRACTICAL Consortium

Christopher A. Haiman[1], Fredrick R. Schumacher[2,3], Sara Benlloch[4,5], Ali Amin Al Olama[6,7], Sonja I. Berndt[8], David V. Conti[1], Fredrik Wiklund[9], Stephen Chanock[8], Susan M. Gapstur[10], Victoria L. Stevens[10], Jyotsna Batra[11,12], Judith Clements[11,12], APCB BioResource[13,14], Henrik Grönberg[15], Nora Pashayan[16,17], Johanna Schleutker[18,19], Demetrius Albanes[8], Stephanie Weinstein[8], Alicja Wolk[20,21], Catharine West[22], Lorelei Mucci[23], Stella Koutros[8], Karina Dalsgaard Sørensen[24,25], Eli Marie Grindedal[26], David E. Neal[27,28,29], Freddie C. Hamdy[30,31], Jenny L. Donovan[32], Ruth C. Travis[33], Robert J. Hamilton[34,35], Sue Ann Ingles[36], Barry S. Rosenstein[37,38], Yong-Jie Lu[39], Graham G. Giles[40,41,42], Ana Vega[43,44,45], Manolis Kogevinas[46,47,48,49], Kathryn L. Penney[50], Janet L. Stanford[51,52], Cezary Cybulski[53], Børge G. Nordestgaard[54,55], Hermann Brenner[56,57,58], Christiane Maier[59], Jeri Kim[60], Manuel R. Teixeira[61,62], Susan L. Neuhausen[63], Kim De Ruyck[64], Azad Razack[65], Lisa F. Newcomb[51,66], Davor Lessel[67], Radka Kaneva[68], Nawaid Usmani[69,70], Frank Claessens[71], Paul A. Townsend[72], Manuela Gago-Dominguez[73,74], Monique J. Roobol[75], Kay-Tee Khaw[76], Lisa Cannon-Albright[77,78], Hardev Pandha[79], Stephen N. Thibodeau[80], Peter Kraft[81], Elio Riboli[82]

[1]Center for Genetic Epidemiology, Department of Preventive Medicine, Keck School of Medicine, University of Southern California/Norris Comprehensive Cancer Center, Los Angeles, CA 90015, USA

[2]Department of Population and Quantitative Health Sciences, Case Western Reserve University, Cleveland, OH 44106-7219, USA

[3]Seidman Cancer Center, University Hospitals, Cleveland, OH 44106, USA.

[4]Centre for Cancer Genetic Epidemiology, Department of Public Health and Primary Care, University of Cambridge, Strangeways Research Laboratory, Cambridge CB1 8RN, UK

[5]The Institute of Cancer Research, London, SM2 5NG, UK

[6]Centre for Cancer Genetic Epidemiology, Department of Public Health and Primary Care, University of Cambridge, Strangeways Research Laboratory, Cambridge, UK

[7]University of Cambridge, Department of Clinical Neurosciences, Stroke Research Group, R3, Box 83, Cambridge Biomedical Campus, Cambridge CB2 0QQ, UK

[8]Division of Cancer Epidemiology and Genetics, National Cancer Institute, NIH, Bethesda, Maryland, 20892, USA

[9]Department of Medical Epidemiology and Biostatistics, Karolinska Institute, SE-171 77 Stockholm, Sweden

[10]Behavioral and Epidemiology Research Group, Research Program, American Cancer

Society, 250 Williams Street, Atlanta, GA 30303, USA
[11]Australian Prostate Cancer Research Centre-Qld, Institute of Health and Biomedical Innovation and School of Biomedical Sciences, Queensland University of Technology, Brisbane QLD 4059, Australia
[12]Translational Research Institute, Brisbane, Queensland 4102, Australia
[13]Australian Prostate Cancer Research Centre-Qld, Queensland University of Technology, Brisbane; Prostate Cancer Research Program, Monash University, Melbourne; Dame Roma Mitchell Cancer Centre, University of Adelaide, Adelaide; Chris O'Brien Lifehouse and
[14]Translational Research Institute, Brisbane, Queensland, Australia
[15]Department of Medical Epidemiology and Biostatistics, Karolinska Institute, Stockholm, Sweden
[16]Department of Applied Health Research, University College London, London, WC1E 7HB, UK
[17]Centre for Cancer Genetic Epidemiology, Department of Oncology, University of Cambridge, Strangeways Laboratory, Worts Causeway, Cambridge, CB1 8RN, UK
[18]Institute of Biomedicine, Kiinamyllynkatu 10, FI-20014 University of Turku, Finland
[19]Department of Medical Genetics, Genomics, Laboratory Division, Turku University Hospital, PO Box 52, 20521 Turku, Finland
[20]Division of Nutritional Epidemiology, Institute of Environmental Medicine, Karolinska Institutet, SE-171 77 Stockholm, Sweden
[21]Department of Surgical Sciences, Uppsala University, 75185 Uppsala, Sweden
[22]Division of Cancer Sciences, University of Manchester, Manchester Academic Health Science Centre, Radiotherapy Related Research, The Christie Hospital NHS Foundation Trust, Manchester, M13 9PL UK
[23]Department of Epidemiology,Harvard T. H. Chan School of Public Health, Boston, MA 02115, USA
[24]Department of Molecular Medicine, Aarhus University Hospital, Palle Juul-Jensen Boulevard 99, 8200 Aarhus N, Denmark
[25]Department of Clinical Medicine, Aarhus University, DK-8200 Aarhus N
[26]Department of Medical Genetics, Oslo University Hospital, 0424 Oslo, Norway
[27]Nuffield Department of Surgical Sciences, University of Oxford, Room 6603, Level 6, John Radcliffe Hospital, Headley Way, Headington, Oxford, OX3 9DU, UK
[28]University of Cambridge, Department of Oncology, Box 279, Addenbrooke's Hospital, Hills Road, Cambridge CB2 0QQ, UK
[29]Cancer Research UK, Cambridge Research Institute, Li Ka Shing Centre, Cambridge UK

[30]Nuffield Department of Surgical Sciences, University of Oxford, Oxford, OX1 2JD, UK
[31]Faculty of Medical Science, University of Oxford, John Radcliffe Hospital, Oxford, UK
[32]Population Health Sciences, Bristol Medical School, University of Bristol, BS8 2PS, UK
[33]Cancer Epidemiology Unit, Nuffield Department of Population Health, University of Oxford, Oxford, OX3 7LF, UK
[34]Dept. of Surgical Oncology, Princess Margaret Cancer Centre, Toronto ON M5G 2M9, Canada
[35]Dept. of Surgery (Urology), University of Toronto, Canada
[36]Department of Preventive Medicine, Keck School of Medicine, University of Southern California/Norris Comprehensive Cancer Center, Los Angeles, CA 90015, USA
[37]Department of Radiation Oncology and Department of Genetics and Genomic Sciences, Box 1236, Icahn School of Medicine at Mount Sinai, One Gustave L. Levy Place, New York, NY 10029, USA
[38]Department of Genetics and Genomic Sciences, Icahn School of Medicine at Mount Sinai, New York, NY 10029-5674 , USA.
[39]Centre for Molecular Oncology, Barts Cancer Institute, Queen Mary University of London, John Vane Science Centre, Charterhouse Square, London, EC1M 6BQ, UK
[40]Cancer Epidemiology Division, Cancer Council Victoria, 615 St Kilda Road, Melbourne, VIC 3004, Australia
[41]Centre for Epidemiology and Biostatistics, Melbourne School of Population and Global Health, The University of Melbourne, Grattan Street, Parkville, VIC 3010, Australia
[42]Precision Medicine, School of Clinical Sciences at Monash Health, Monash University, Clayton, Victoria 3168, Australia
[43]Fundación Pública Galega Medicina Xenómica, Santiago De Compostela, 15706, Spain.
[44]Instituto de Investigación Sanitaria de Santiago de Compostela, Santiago De Compostela, 15706, Spain.
[45]Centro de Investigación en Red de Enfermedades Raras (CIBERER), Spain
[46]ISGlobal, Barcelona, Spain
[47]IMIM (Hospital del Mar Medical Research Institute), Barcelona, Spain
[48]Universitat Pompeu Fabra (UPF), Barcelona, Spain
[49]CIBER Epidemiología y Salud Pública (CIBERESP), Madrid, Spain
[50]Channing Division of Network Medicine, Department of Medicine, Brigham and Women's Hospital/Harvard Medical School, Boston, MA 02184, USA
[51]Division of Public Health Sciences, Fred Hutchinson Cancer Research Center, Seattle,

Washington, 98109-1024, USA
[52]Department of Epidemiology, School of Public Health, University of Washington, Seattle, Washington 98195, USA
[53]International Hereditary Cancer Center, Department of Genetics and Pathology, Pomeranian Medical University, Szczecin, Poland
[54]Faculty of Health and Medical Sciences, University of Copenhagen, 2200 Copenhagen, Denmark
[55]Department of Clinical Biochemistry, Herlev and Gentofte Hospital, Copenhagen University Hospital, Herlev, 2200 Copenhagen, Denmark
[56]Division of Clinical Epidemiology and Aging Research, German Cancer Research Center (DKFZ), D-69120, Heidelberg, Germany
[57]German Cancer Consortium (DKTK), German Cancer Research Center (DKFZ), D-69120 Heidelberg, Germany
[58]Division of Preventive Oncology, German Cancer Research Center (DKFZ) and National Center for Tumor Diseases (NCT), Im Neuenheimer Feld 460
69120 Heidelberg, Germany
[59]Humangenetik Tuebingen, Paul-Ehrlich-Str 23, D-72076 Tuebingen, Germany
[60]The University of Texas M. D. Anderson Cancer Center, Department of Genitourinary Medical Oncology, 1515 Holcombe Blvd., Houston, TX 77030, USA
[61]Department of Genetics, Portuguese Oncology Institute of Porto (IPO-Porto), Porto, Portugal
[62]Biomedical Sciences Institute (ICBAS), University of Porto, Porto, Portugal
[63]Department of Population Sciences, Beckman Research Institute of the City of Hope, 1500 East Duarte Road, Duarte, CA 91010, 626-256-HOPE (4673)
[64]Ghent University, Faculty of Medicine and Health Sciences, Basic Medical Sciences, Proeftuinstraat 86, B-9000 Gent
[65]Department of Surgery, Faculty of Medicine, University of Malaya, 50603 Kuala Lumpur, Malaysia
[66]Department of Urology, University of Washington, 1959 NE Pacific Street, Box 356510, Seattle, WA 98195, USA
[67]Institute of Human Genetics, University Medical Center Hamburg-Eppendorf, D-20246 Hamburg, Germany
[68]Molecular Medicine Center, Department of Medical Chemistry and Biochemistry, Medical University of Sofia, Sofia, 2 Zdrave Str., 1431 Sofia, Bulgaria
[69]Department of Oncology, Cross Cancer Institute, University of Alberta, 11560 University

Avenue, Edmonton, Alberta, Canada T6G 1Z2
[70]Division of Radiation Oncology, Cross Cancer Institute, 11560 University Avenue, Edmonton, Alberta, Canada T6G 1Z2
[71]Molecular Endocrinology Laboratory, Department of Cellular and Molecular Medicine, KU Leuven, BE-3000, Belgium
[72]Division of Cancer Sciences, Manchester Cancer Research Centre, Faculty of Biology, Medicine and Health, Manchester Academic Health Science Centre, NIHR Manchester Biomedical Research Centre, Health Innovation Manchester, Univeristy of Manchester, M13 9WL
[73]Genomic Medicine Group, Galician Foundation of Genomic Medicine, Instituto de Investigacion Sanitaria de Santiago de Compostela (IDIS), Complejo Hospitalario Universitario de Santiago, Servicio Galego de Saúde, SERGAS, 15706, Santiago de Compostela, Spai
[74]University of California San Diego, Moores Cancer Center, La Jolla, CA 92037, USA
[75]Department of Urology, Erasmus University Medical Center, 3015 CE Rotterdam, The Netherlands
[76]Clinical Gerontology Unit, University of Cambridge, Cambridge, CB2 2QQ, UK
[77]Division of Epidemiology, Department of Internal Medicine, University of Utah School of Medicine, Salt Lake City, Utah, USA
[78]George E. Wahlen Department of Veterans Affairs Medical Center, Salt Lake City, Utah 84148, USA
[79]The University of Surrey, Guildford, Surrey, GU2 7XH
[80]Department of Laboratory Medicine and Pathology, Mayo Clinic, Rochester, MN 55905, USA
[81]Program in Genetic Epidemiology and Statistical Genetics, Department of Epidemiology, Harvard School of Public Health, Boston, MA, USA
[82]Department of Epidemiology and Biostatistics, School of Public Health, Imperial College London, SW7 2AZ, UK

### Appendix 2. Funding sources for the PRACTICAL consortium


CRUK and PRACTICAL consortium

This work was supported by the Canadian Institutes of Health Research, European Commission's Seventh Framework Programme grant agreement n° 223175 (HEALTH-F2-2009-223175), Cancer Research UK Grants C5047/A7357, C1287/A10118, C1287/A16563, C5047/A3354, C5047/A10692, C16913/A6135, and The National Institute of Health (NIH) Cancer Post-Cancer GWAS initiative grant: No. 1 U19 CA 148537-01 (the GAME-ON initiative).

We would also like to thank the following for funding support: The Institute of Cancer Research and The Everyman Campaign, The Prostate Cancer Research Foundation, Prostate Research Campaign UK (now Prostate Action), The Orchid Cancer Appeal, The National Cancer Research Network UK, The National Cancer Research Institute (NCRI) UK. We are grateful for support of NIHR funding to the NIHR Biomedical Research Centre at The Institute of Cancer Research and The Royal Marsden NHS Foundation Trust.

The Prostate Cancer Program of Cancer Council Victoria also acknowledge grant support from The National Health and Medical Research Council, Australia (126402, 209057, 251533, , 396414, 450104, 504700, 504702, 504715, 623204, 940394, 614296,), VicHealth, Cancer Council Victoria, The Prostate Cancer Foundation of Australia, The Whitten Foundation, PricewaterhouseCoopers, and Tattersall's. EAO, DMK, and EMK acknowledge the Intramural Program of the National Human Genome Research Institute for their support.

Genotyping of the OncoArray was funded by the US National Institutes of Health (NIH) [U19 CA 148537 for ELucidating Loci Involved in Prostate cancer SuscEptibility (ELLIPSE) project and X01HG007492 to the Center for Inherited Disease Research (CIDR) under contract number HHSN268201200008I].

This study would not have been possible without the contributions of the following: Coordination team, bioinformatician and genotyping centers: Genotyping at CCGE, Cambridge: Caroline Baines and Don Conroy

Funding for the iCOGS infrastructure came from: the European Community's Seventh Framework Programme under grant agreement n° 223175 (HEALTH-F2-2009-223175) (COGS), Cancer Research UK (C1287/A10118, C1287/A 10710, C12292/A11174,

C1281/A12014, C5047/A8384, C5047/A15007, C5047/A10692, C8197/A16565), the National Institutes of Health (CA128978) and Post-Cancer GWAS initiative (1U19 CA148537, 1U19 CA148065 and 1U19 CA148112 - the GAME-ON initiative), the Department of Defence (W81XWH-10-1-0341), the Canadian Institutes of Health Research (CIHR) for the CIHR Team in Familial Risks of Breast Cancer, Komen Foundation for the Cure, the Breast Cancer Research Foundation, and the Ovarian Cancer Research Fund.

This study would not have been possible without the contributions of the following: Per Hall (COGS); Douglas F. Easton, Paul Pharoah, Kyriaki Michailidou, Manjeet K. Bolla, Qin Wang (BCAC), Andrew Berchuck (OCAC), Rosalind A. Eeles, Douglas F. Easton, Ali Amin Al Olama, Zsofia Kote-Jarai, Sara Benlloch (PRACTICAL), Georgia Chenevix-Trench, Antonis Antoniou, Lesley McGuffog, Fergus Couch and Ken Offit (CIMBA), Joe Dennis, Alison M. Dunning, Andrew Lee, and Ed Dicks, Craig Luccarini and the staff of the Centre for Genetic Epidemiology Laboratory, Javier Benitez, Anna Gonzalez-Neira and the staff of the CNIO genotyping unit, Jacques Simard and Daniel C. Tessier, Francois Bacot, Daniel Vincent, Sylvie LaBoissière and Frederic Robidoux and the staff of the McGill University and Génome Québec Innovation Centre, Stig E. Bojesen, Sune F. Nielsen, Borge G. Nordestgaard, and the staff of the Copenhagen DNA laboratory, and Julie M. Cunningham, Sharon A. Windebank, Christopher A. Hilker, Jeffrey Meyer and the staff of Mayo Clinic Genotyping Core Facility

Additional funding and acknowledgments from studies in PRACTICAL:

Information of the PRACTICAL consortium can be found at http://practical.icr.ac.uk/

BioVU

The dataset(s) used for the analyses described were obtained from Vanderbilt University Medical Center's BioVU, which is supported by institutional funding and by the National Center for Research Resources, Grant UL1 RR024975-01 (which is now at the National Center for Advancing Translational Sciences, Grant 2 UL1 TR000445-06).

CPDR

Uniformed Services University for the Health Sciences HU0001-10-2-0002 (PI: David G. McLeod, MD)

EPICAP

The EPICAP study was supported by grants from Ligue Nationale Contre le Cancer; Institut National du Cancer (INCa); Fondation ARC; Fondation de France; Agence Nationale de sécurité sanitaire de l'alimentation, de l'environnement et du travail (ANSES); Ligue départementale du Val de Marne. The EPICAP study group would like to thank all urologists, Antoinette Anger and Hasina Randrianasolo (study monitors), Anne-Laure Astolfi, Coline Bernard, Oriane Noyer, Marie-Hélène De Campo, Sandrine Margaroline, Louise N'Diaye, Sabine Perrier-Bonnet (Clinical Research nurses)

KARUPROSTATE

The Karuprostate study was supported by the the Frech National Health Directorate, the Association pour la Recherche sur le Cancer, la Ligue Nationale contre le Cancer, the French Agency for Environmental and Occupational Health Safety (ANSES) and by the Association pour la Recherche sur les Tumeurs de la Prostate. We would like to thank Séverine Ferdinand for valuable contributions to the study.

MOFFITT

The Moffitt group was supported by the US National Cancer Institute (R01CA128813, PI: J.Y. Park).

NMHS

Funding for the Nashville Men's Health Study (NMHS) was provided by the National Institutes of Health Grant numbers: RO1CA121060

PCaP

The North Carolina - Louisiana Prostate Cancer Project (PCaP) and the Health Care Access and Prostate Cancer Treatment in North Carolina (HCaP-NC) study are carried out as collaborative studies supported by the Department of Defense contract DAMD 17-03-2-0052 and the American Cancer Society award RSGT-08-008-01-CPHPS, respectively.

The authors thank the staff, advisory committees and research subjects participating in the PCaP and HCaP-NC studies for their important contributions.

PROtEuS

PROtEuS was supported financially through grants from the Canadian Cancer Society [13149, 19500, 19864, 19865] and the Cancer Research Society, in partnership with the Ministère de l'enseignement supérieur, de la recherche, de la science et de la technologie du Québec, and the Fonds de la recherche du Québec - Santé.PROtEuS would like to thank its collaborators and research personnel, and the urologists involved in subjects recruitment. We also wish to acknowledge the special contribution made by Ann Hsing and Anand Chokkalingam to the conception of the genetic component of PROtEuS.

SABOR

The SABOR research is supported by NIH/NCI Early Detection Research Network, grant U01 CA0866402-18. Also supported by the Cancer Center Support Grant to the Mays Cancer Center from the National Cancer Institute (US) P30 CA054174

SCCS

SCCS is funded by NIH grant R01 CA092447, and SCCS sample preparation was conducted at the Epidemiology Biospecimen Core Lab that is supported in part by the Vanderbilt-Ingram Cancer Center (P30 CA68485). Data on SCCS cancer cases used in this publication were provided by the Alabama Statewide Cancer Registry; Kentucky Cancer Registry, Lexington, KY; Tennessee Department of Health, Office of Cancer Surveillance; Florida Cancer Data System; North Carolina Central Cancer Registry, North Carolina Division of Public Health; Georgia Comprehensive Cancer Registry; Louisiana Tumor Registry; Mississippi Cancer Registry; South Carolina Central Cancer Registry; Virginia Department of Health, Virginia Cancer Registry; Arkansas Department of Health, Cancer Registry, 4815 W. Markham, Little Rock, AR 72205. The Arkansas Central Cancer Registry is fully funded by a grant from National Program of Cancer Registries, Centers for Disease Control and Prevention (CDC). Data on SCCS cancer cases from Mississippi were collected by the Mississippi Cancer Registry which participates in the National Program of Cancer Registries (NPCR) of the Centers for Disease Control and Prevention (CDC). The contents of this publication are solely the responsibility of the authors and do not necessarily represent the official views of the CDC or the Mississippi Cancer Registry.

SCPCS

SCPCS is funded by CDC grant S1135-19/19, and SCPCS sample preparation was conducted at the Epidemiology Biospecimen Core Lab that is supported in part by the Vanderbilt-Ingram Cancer Center (P30 CA68485).

SFPCS

SFPCS was funded by California Cancer Research Fund grant 99-00527V-10182

SWOG-PCPT / SWOG-SELECT

PCPT and SELECT are funded by Public Health Service grants U10CA37429 and 5UM1CA182883 from the National Cancer Institute. The authors thank the site investigators and staff and, most importantly, the participants from PCPT who donated their time to this trial.

UKGPCS

UKGPCS would also like to thank the following for funding support: The Institute of Cancer Research and The Everyman Campaign, The Prostate Cancer Research Foundation, Prostate Research Campaign UK (now Prostate Action), The Orchid Cancer Appeal, The National Cancer Research Network UK, The National Cancer Research Institute (NCRI) UK. We are grateful for support of NIHR funding to the NIHR Biomedical Research Centre at The Institute of Cancer Research and The Royal Marsden NHS Foundation Trust. UKGPCS should also like to acknowledge the NCRN nurses, data managers and Consultants for their work in the UKGPCS study.UKGPCS would like to thank all urologists and other persons involved in the planning, coordination, and data collection of the study. KM and AL were in part supported from the NIHR Manchester Biomedical Research Centre

WUGS

WUGS would like to thank the following for funding support: The Anthony DeNovi Fund, the Donald C. McGraw Foundation, and the St. Louis Men’s Group Against Cancer.
